# Cerebrospinal fluid CXCL13 identifies a subgroup of people living with HIV with prominent intrathecal synthesis, immune activation, and neurocognitive impairment regardless of effective antiretroviral therapy

**DOI:** 10.1101/2022.12.13.22283427

**Authors:** Mattia Trunfio, Lorenzo Mighetto, Laura Napoli, Cristiana Atzori, Marco Nigra, Giulia Guastamacchia, Stefano Bonora, Giovanni Di Perri, Andrea Calcagno

## Abstract

**Background:** Plasma C-X-C-motif chemokine ligand-13 (CXCL13) has been linked to disease progression and mortality in people living with HIV (PLWH) and is a candidate target for immune-based strategies for HIV cure. Its role in central nervous system (CNS) of PLWH has not been detailed. We described CSF CXCL13 levels and its potential associations with neurological outcomes.

**Methods:** Cross-sectional study enrolling PLWH without confounding for CXCL13 production. Subjects were divided according to CSF HIV-RNA in controllers (<20 cp/mL) and viremics. CSF CXCL13, and biomarkers of blood-brain barrier (BBB) impairment, intrathecal synthesis, and immune activation were measured by commercial immunoturbidimetric and ELISA assays. All subjects underwent neurocognitive assessment. Sensitivity analyses were conducted in subjects with intact BBB only.

**Results:** 175 subjects were included. Prevalence of detectable CSF CXCL13 was higher in viremics (31.4%) compared to controllers (13.5%; OR 2.9 [1.4-6.3], p=0.006), but median CSF levels did not change (15.8 [8.2-91.0] vs 10.0 [8.1-14.2] pg/mL). In viremics (n=86), CXCL13 associated with higher CSF HIV-RNA, proteins, neopterin, Tourtelotte index, and CSF-to-serum albumin ratio. In controllers (n=89), CXCL13 associated with higher CD4+T-cells count, CD4/CD8 ratio, CSF proteins, neopterin, and several intrathecal synthesis markers. Detection of CSF CXCL13 in controllers increased the likelihood of HIV-associated neurocognitive disorders (58.3% vs 28.6%, p=0.041) and HIV-related CNS disorders (8.3% vs 0%, p=0.011). Sensitivity analyses confirmed all these findings.

**Conclusions:** CSF CXCL13 identified a subgroup of PLWH presenting increased CNS IgG synthesis, and immune activation. In controllers, CSF CXCL13 associated with increased likelihood of neurocognitive impairment and HIV-related CNS disorders.

**Graphical abstract:** 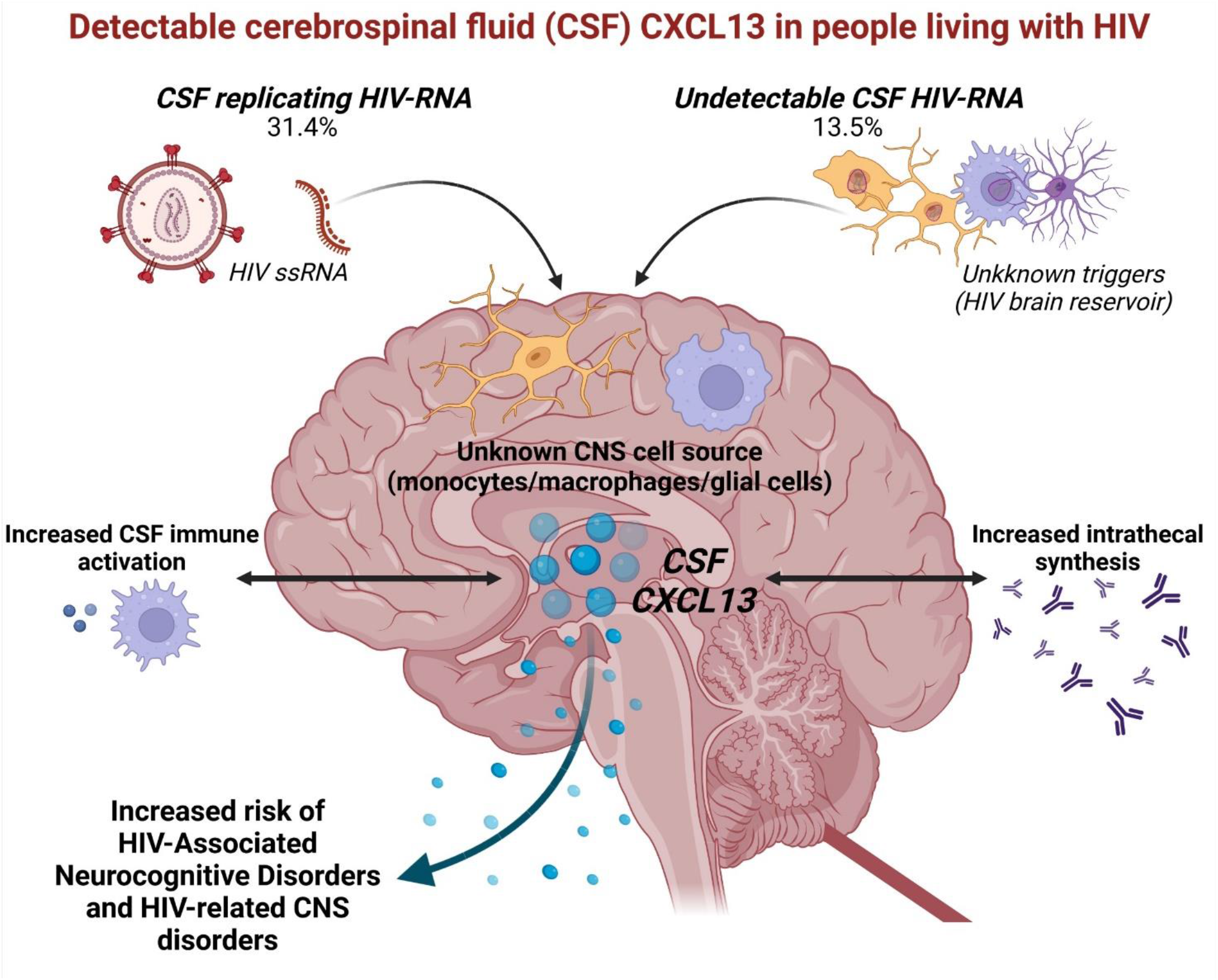

## Introduction

Despite the success of combination antiretroviral therapy (cART), people living with HIV (PLWH) still suffer from chronic inflammation and persistent immune activation that eventually can lead to increased risk of mortality and development of non-AIDS events (Boulougoura and Sereti 2016; Sauce et al. 2021; Looby et al. 2022). One of the main driver of such events is represented by the persistence of HIV in cellular and tissue reservoirs, where ongoing viral antigen low-level stimulation takes place with or without residual replication (Ploquin et al. 2016; Avettand-Fènoël et al. 2016). Therefore, therapeutic strategies to tackle residual immune activation through direct or indirect actions on viral reservoires are needed to improve the life expectancy and the quality of life of PLWH (Siliciano and Siliciano 2014; Rajasuriar et al. 2015; Hsu and Ananworanich 2018; Board et al. 2021). Indeed, secondary lymphoid tissues and lymphoid/myeloid cells within them are one of the most important contributors of the whole viral reservoir (Churchill et al. 2016; Cohn et al. 2020). With this perspective, several biomarkers of immune activation/inflammation have been validated, but to date, no single biomarker could account for both lymphoid and myeloid cells activation. Furthermore, humoral immune dysfunction in HIV infection has been partially neglected although HIV-associated B cells perturbations contribute to fuelling immune activation (Moir and Fauci 2014; de Bree and Lynch 2016).

C-X-C motif chemokine ligand 13 (CXCL13) is expressed in all primary, secondary and tertiary lymphoid tissues (such as liver, spleen, lymph nodes, and gut and ectopic lymphoid tissues that develop at sites of chronic inflammation in nonlymphoid organs) by T follicular helper cells (Tfh), follicular dendritic cells, stromal cells, monocytes and macrophages (Legler et al. 1998; Gunn et al. 1998; Carlsen et al. 2004; Cohen-Kaminsky et al. 2014). CXCL13 is the ligand of the 7 transmembrane-domain G protein-coupled chemokine receptor CXCR5, which is expressed by B cells, follicular NK and CD8+ cytotoxic T cells, Tfh, T regulatory, and dendritic cells (Legler et al. 1998; Gunn et al. 1998; Carlsen et al. 2004; Bekele Feyissa et al. 2021). Its main role is regulating the lymphoid tissue development by attracting lymphoid tissue inducer cells during the lymphoid tissue development (Legler et al. 1998; Gunn et al. 1998; Carlsen et al. 2004; Bekele Feyissa et al. 2021). CXCL13 also attracts B cells to these sites and is fundamental for compartmentalization of germinal centers, promoting the localization of Tfh and B cells in the light zone, where several maturation processes for the functioning of these cells occur (Legler et al. 1998; Gunn et al. 1998; Carlsen et al. 2004; Bekele Feyissa et al. 2021). Not surprisingly, this chemokine has been associated with multiple biological processes such as germinal centers activity after infections and vaccination (Masouris et al. 2020; Bekele Feyissa et al. 2021) with eventual antibodies production (Havenar-Daughton et al. 2016; Mabuka et al. 2017; Paris et al. 2017; Dugast et al. 2017), recruitment of cells to inflammatory and cancer sites (Cohen-Kaminsky et al. 2014; Kazanietz et al. 2019), and also with autoimmune diseases (Pan et al. 2022) and hematologic cancers (Hussain et al. 2013). Due to its plethora of activities and effects upon several cell types, CXCL13 can be regarded as a biomarker of both lymphoid (humoral and cellular) and myeloid cell activation and networking.

In the setting of HIV infection there is growing interest in CXCL13; this chemokine has been linked to several HIV-related processes (under investigation for tailored immune-based treatments) and it has been involved in predicting vaccine responses (including anti-HIV vaccination in HIV cure strategies) (Havenar-Daughton et al. 2016; Mabuka et al. 2017; Dugast et al. 2017; Bekele Feyissa et al. 2021; Guo et al. 2022). Compared to healthy HIV-negative controls and HIV-positive elite controllers, plasma levels of CXCL13 have been reported higher in HIV acute and chronic stages, regardless of cART (Cagigi et al. 2008; Regidor et al. 2011; Mehraj et al. 2019), and they have been correlated with disease progression, mortality, microbial translocation, myeloid and lymphoid cell activation, and with AIDS-associated B-cells lymphoma development (Hussain et al. 2013; Wada et al. 2016; Mehraj et al. 2019; Bekele Feyissa et al. 2021). Despite Tfh seem to represent the largest contributor to CXCL13 production, it has been also demonstrated an induction in CXCL13 production directly made by HIV-1 single strand RNA in monocytes (Cohen et al. 2015).

As for vaccine responses, higher plasma CXCL13 levels were detected among non-responders compared to responders to anti-HBV vaccine with an inverse correlation between CXCL13 and anti-HBs levels (Paris et al. 2017), suggesting that chronic hyper-activation of germincal centers could impair vaccine responses in PLWH. Furthermore, plasma CXCL13 levels have been correlated with anti-HIV broadly neutralizing antibodies development (Havenar-Daughton et al. 2016; Dugast et al. 2017) and with the expansion of a subset of HIV-specific CD4+ T-helper 1 cells represented by circulating Tfh, involved in immunological memory (Niessl et al. 2020). Overall, CXCL13 could have a double-faced role: it is involved in potentially protective HIV-specific immunity development, but it could also drive the hyperinflammatory state, and could contribute to hypergammaglobulinemia, to the perturbations in humoral responses, and to the replenishment of HIV reservoir through the increase in follicular T cells survival and accumulation in lymphoid follicles, the major sanctuary for HIV persistence (Bekele Feyissa et al. 2021).

Immunological responses in different body sites, as well as the HIV reservoirs and immunological sanctuaries, can be compartmentalized (Beck et al. 2016; de Almeida et al. 2016; Stefic et al. 2017). To date, only two studies reported cerebrospinal fluid (CSF) levels of CXCL13 in neurologically healthy PLWH, but HIV infection was used as control group for neuroborreliosis and other central nervous system (CNS) infections and no description of the CXCL13/CXCR5 axis in the CNS of PLWH has been detailed (van Burgel et al. 2011; Bremell et al. 2013). Despite the absence of germinal centers, CNS possess lymphoid cells and monocytes that are able to produce CXCL13 (Wang and van de Pavert 2022) together with several cell lines able to silently harbour HIV as cellular reservoir (Wallet et al. 2019). Being CNS an immunological sanctuary, the amount and type of humoral responses within CNS can differ from those that take place in periphery (Beck et al. 2016; de Almeida et al. 2016; Stefic et al. 2017). Therefore, the absence of knowledge about CXCL13/CXCR5 axis within the CNS of PLWH could represent a relevant limitation in the use of this chemokine as therapeutic target or in predictive models for eradication strategies. Furthermore, the assessment of CXCL13 in CSF could inform on several chronic mechanisms underlying the compartmentalized intrathecal synthesis of HIV-specific and -unspecific immunoglobulins. Indeed, despite persistent intrathecal synthesis has been repeatedly observed and associated with clinical and biological unfavourable outcomes, such as blood-brain barrier (BBB) and neurocognitive impairment, its drivers and the antigen targets remain poorly understood in PLWH (Bonnan et al. 2015).

In the light of this knowledge gap aim of this study was to measure CSF CXCL13 levels in neurologically symptomatic and asymptomatic adult PLWH, both cART-naive and cART-treated, and to describe whether CSF CXCL13 associates with other better known CSF biomarkers of inflammation, immune activation, blood-brain barrier (BBB) integrity, and intrathecal synthesis, as well as with viro-immunological parameters.

## Materials and Methods

We performed a retrospective cross-sectional study nested in ongoing prospective studies on CSF of HIV-positive patients at our centre, Infectious Diseases Unit, Amedeo di Savoia hospital (Torino, Italy). We enrolled cART-naïve and cART-treated adult patients undergoing lumbar puncture (LP) for clinical or research reasons from January 2010 to January 2020. Inclusion criteria were: age≥18 years, western-blot confirmed HIV infection, signed written informed consent and the availability of at least 1 mL of stored CSF to allow for CXCL13 measurement. Exclusion criteria were chosen to rule out potential conditions affecting CXCL13 production: LP contraindications, untreated infections by *spirochaetes*, lymphoproliferative disorders with or without CNS involvement, infectious, autoimmune and/or inflammatory CNS disorders, CNS primary or metastatic cancers, and CSF low-level replicating EBV and CMV.

The primary objective was to describe CSF CXCL13 concentrations among naïve and on cART HIV-positive adult patients without CNS confounding conditions. Secondary objective were: 1) to assess potential associations of demographic, clinical, and of viro-immunological parameters with CSF CXCL13 levels; 2) to assess potential associations of CSF biomarkers of immune activation, BBB impairment, inflammation and intrathecal synthesis with CSF CXCL13 levels.

Considering CSF HIV-RNA as relevant collider for the relationship between CSF CXCL13 and several of the assessed variables (Cohen et al. 2015; Bonnan et al. 2015; Mehraj et al. 2019), the study population was divided according to the presence (CSF HIV-RNA≥20 cp/mL, CSF viremic) or absence (<20 cp/mL, CSF controllers) of replicating HIV within CSF. Furthermore, to reduce potential quantification bias due to passive leakage of plasma CXCL13 (not measured) into CSF, sensitivity analyses were performed in the study population after removing participants with impaired BBB. This was deemed also as a confirmation of the intrathecal origin of the measured immunoglobulin synthesis that was already normalized for BBB permeability.

CXCL13 was measured in CSF samples stored at -80°C and collected between January 2010 to June 2019. The chemokine was measured by standard quantitative sandwich enzyme-linked immunosorbent assay technology (Quantikine ELISA assay; R&D Systems Europe, Ltd. Abingdon, UK). The kit was validated for human fluids and tissue homogenates, including CSF. The assay sensitivity has been estimated by manufacturer to be 1,64 pg/mL and the detection range to be 3,5-10.000 pg/ml. Therefore, all CSF samples with less than 3,5 pg/mL were classified as negative for CXCL13. In terms of assay specificity, the manufacturer tested the assay with several human cytokines and chemokines (CXCL16, ENA-78, GCP-2, GROα, GROβ, GROγ, IL-8, IL-8 endothelial cell-derived, IP-10, I-TAC, MIG, NAP-2, SDF-1α, SDF-1β); no significant cross-reactivity nor interference was observed.

The remaining CSF analyses were performed immediately after LP collection and consisted of cells, proteins and glucose assessment, HIV-1 RNA quantification by Roche Amplicor assay v2.0 (Hoffman-La Roche, Basel, Switzerland) with a limit of quantification of 20 copies/mL and of the quantitative determination of albumin and immunoglobulin (contemporarily in CSF and serum) by immunoturbidimetric methods (AU 5800, Beckman Coulter, Brea, CA, USA). Reibergram was used to calculate Tourtelotte and Tibbling Index, CSF-to-Serum Albumin ratio (CSAR), IgG Index and the proportion of intrathecal origin of immunoglobulins over the whole amount of CSF immunoglobulins detected (CSF % synthesis), as previously described (Reiber 1995). Reference values were as follows: CSAR <6.5 in subjects of up to 40 years and <8 in patients aged 41 years and above; IgG index <0.7, intrathecal synthesis 0.0%. CSF neopterin and S100β were measured by validated ELISA methods (DRG Diagnostics, Marburg, Germany and DIAMETRA Srl, Spello, Italy, respectively). Reference values were as follows: neopterin <1.5 ng/ml and S100β <380 pg/ml.

Demographic, clinical data, and viro-immunological parameters were collected from clinical records. All the subjects underwent neurocognitive assessment (Trail Making test part A and B, Stroop colour test, Digit Span forward and backward, Digit Symbol, Corsi and Disyllabic Words Serial Repetition tests, Free and Cued Selective Reminding test, Story Recall, Frontal Assessment battery, immediate and delayed recall of Rey-Osterrieth complex figure, phonemic, semantic and alternate verbal fluency and Grooved Pegboard for dominant and non-dominant hand) and 1-Tesla brain magnetic resonance imaging. HIV-associated neurocognitive disorders (HAND) were diagnosed according to Frascati’s criteria, as previously detailed (Antinori et al. 2007). At the end of the neurological assessment, the subjects were classified into the following categories: a) asymptomatic: in case of lumbar puncture for research purposes and negative findings at the assessment; b) neurological complaints: in case of reported or objectified neurological signs or symptoms in the absence of CSF, MRI and neurocognitive alterations; c) HAND: in case of altered neurocognitive results compatible with HAND diagnosis and no other alteration found at the end of the assessment; d) HIV-related disorders: in case of symptomatic and asymptomatic CSF viral escape (as previously defined)(Trunfio et al. 2019) or HIV meningo-encephalitis.

Continuous variables were reported as medians (interquartile range) and discrete variables were reported as absolute number (proportion). Appropriate tests (Mann–Whitney test, Kruskal-Wallis test, Chi-square test, Fisher exact test and Pearson’s and Spearman’s correlations) were used according to normal and non-normal distribution of the variables assessed by the Shapiro–Wilk statistics. Linear regressions were run to assess co-linearity between CSF CXCL13 levels and variables with significant correlations. The significance threshold to reject the null hypothesis was set at 5% (α=0.05, two-tailed predictions). Data analysis was performed using SPSS v27 (IBM Corp., Armonk, NY, USA).

The work has been carried out in accordance with <u>The Code of Ethics of the World Medical Association</u> (Declaration of Helsinki) for experiments involving humans and approved by the Ethics Committee of San Luigi Hospital, Orbassano (PRODIN study, protocol code 103/2015, approved on 22 June 2015). Informed consent was obtained from each study participant. This work received funding specifically dedicated to the Department of Medical Sciences from the Italian Ministry for Education, University and Research (Ministero dell’Istruzione, dell’Università e della Ricerca-MIUR) under the programme “Dipartimenti di Eccellenza 2018-2022” (project n. D15D18000410001).

## Results

Among the 501 LP performed to 364 HIV-positive subjects during the study period, 163 patients were excluded for meeting at least one exclusion criteria; further 26 subjects had no available stored CSF sample. Eventually, 175 patients were included.

### 1. CSF CXCL13 in CSF viremic subjects

Eighty-six subjects had detectable CSF HIV-RNA: 57 were not on cART (66.3%), 20 had compartmentalized or systemic viral failure (23.2%), and 9 started cART since less than 6 months before LP (10.5%). They were mostly Caucasian (82.6%) male (76.7%), with advanced HIV infection despite the relatively recent diagnosis: median CD4+ T cell count and CD4/CD8 ratio were 82 (35-273) cells/mmc and 0.2 (0.1-0.4). The prevalence of BBB impairment and intrathecal synthesis were 23.2% and 54.6%, respectively (as shown in Tab.1).

**Table 1.** Comparison between CSF viremic subjects with versus without detectable CSF CXCL13: demographic, clinical, viro-immunological and CSF biomarkers. CXCL13, C-X-C motif chemokine ligand 13; MSM, males who have sex with other males; pIVDU, previous intra-venous drug users; PI, protease inhibitor; NRTIs, nucleoside reverse transcriptase inhibitors; nNRTI, non-nucleoside reverse transcriptase inhibitor; INI, integrase strand transfer inhibitor; CSF, cerebrospinal fluid; HAND, HIV-Associated neurocognitive disorders; CNS, central nervous system; CSAR, cerebrospinal fluid-to-serum albumin ratio; BBB, blood-brain barrier; S100b, S100 beta protein.

| Parameter | CSF viremic<br>(n=86) | Detectable CSF<br>CXCL13 (n=27) | Undetectable<br>CSF CXCL13<br>(n=59) | P |
| --- | --- | --- | --- | --- |
| Age, years | 45 (38-50) | 47 (38-52) | 44 (39-50) | 0.473 |
| Male sex, n | 66 (76.7%) | 20 (74.1%) | 46 (78.0%) | 0.693 |
| Caucasian, n | 71 (82.6%) | 24 (88.9%) | 47 (79.7%) | 0.298 |
| Exposure route, n |  |  |  | 0.733 |
| MSM | 27 (31.4%) | 8 (29.6%) | 19 (32.2%) |  |
| Heterosexual | 30 (34.9%) | 11 (40.7%) | 19 (32.2%) |  |
| pIVDU | 29 (33.7%) | 8 (29.6%) | 21 (35.5%) |  |
| On cART, n | 29 (33.7%) | 9 (33.3%) | 20 (33.9%) | 0.959 |
| Regimen, n |  |  |  | 0.498 |
| Dual | 5/29 (17.2%) | 3/9 (33.3%) | 2/20 (10.0%) |  |
| PI-2NRTIs | 7/29 (24.1%) | 1/9 (11.1%) | 6/20 (30.0%) |  |
| nNRTI-2NRTIs | 5/29 (17.2%) | 1/9 (11.1%) | 4/20 (20.0%) |  |
| INI-2NRTIs | 5/59 (17.2%) | 2/9 (22.2%) | 3/20 (15.0%) |  |
| Others | 7/29 (24.1%) | 2/9 (22.2%) | 5/20 (25.0%) |  |
| Plasma HIV-RNA, Log10 cp/mL | 4.99 (2.31-5.83) | 4.94 (1.91-5.63) | 5.18 (2.38-5.94) | 0.829 |
| Undetectable plasma HIV-RNA, n | 15 (17.4%) | 4 (14.8%) | 11 (18.6%) | 0.842 |
| CSF HIV-RNA, Log10 cp/mL | 3.21 (2.14-4.41) | 4.23 (2.19-4.81) | 2.78 (2.11-3.89) | **0.025** |
| CSF escape, n | 17 (19.8%) | 6 (22.2%) | 11 (18.6%) | 0.701 |
| CD4 count, cells/mm<sup>3</sup> | 82 (35-273) | 72 (34-198) | 82 (35-282) | 0.962 |
| CD4/CD8 ratio | 0.2 (0.1-0.4) | 0.2 (0.1-0.3) | 0.1 (0.1-0.5) | 0.417 |
| CD4 nadir, cells/mm<sup>3</sup> | 56 (21-159) | 70 (26-128) | 49 (21-169) | 0.579 |
| Time since HIV diagnosis, months | 17 (1-151) | 31 (1-150) | 15 (1-154) | 0.666 |
| Clinical category, n |  |  |  | 0.959 |
| Neurological complaints | 7 (8.1%) | 2 (7.4%) | 5 (8.4%) | 0.850 |
| HAND | 15 (17.4%) | 4 (14.8%) | 11 (18.6%) | 0.780 |
| Asymptomatic | 38 (44.2%) | 12 (44.4%) | 26 (44.1%) | 0.974 |
| CNS HIV-related | 26 (30.2%) | 9 (33.3%) | 17 (28.8%) | 0.709 |
| Previous syphilis, n | 10 (11.6%) | 1 (3.7%) | 9 (15.2%) | 0.121 |
| CSF cells | 0 (0-1) | 0 (0-3) | 0 (0-0) | 0.837 |
| CSF proteins | 47 (35-58) | 55 (39-65) | 44 (32-55) | **0.011** |
| CSF glucose | 52 (46-59) | 50 (39-58) | 54 (48-60) | 0.070 |
| Tourtelotte | 6.5 (0.1-24.2) | 20.4 (4.6-47.1) | 5.6 (0.0-16.4) | **0.014** |
| Tibbling | 0.8 (0.6-1.2) | 1.0 (0.6-1.2) | 0.8 (0.5-1.1) | 0.246 |
| CSAR | 5.2 (3.6-7.1) | 5.7 (3.8-7.9) | 4.8 (3.4-7.1) | **0.013** |
| Impaired BBB | 20 (23.2%) | 8 (29.6%) | 12 (20.3%) | 0.344 |
| IgG index | 0.45 (0.31-0.70) | 0.60 (0.34-0.98) | 0.40 (0.26-0.62) | 0.182 |
| CSF synthesis, n | 47 (54.6%) | 18 (66.7%) | 29 (49.1%) | 0.130 |
| CSF % synthesis, % | 38.0 (27.0-47.0) | 37.5 (28.5-52.0) | 38.0 (25.5-46.5) | 0.620 |
| Neopterin | 1.5 (1.0-3.7) | 3.42 (0.95-14.11) | 1.32 (0.97-2.69) | **<0.001** |
| S100b | 127.9 (73.0-206.1) | 143.9 (77.6-296.7) | 124.1 (73.0-162.3) | 0.716 |

Among these, 27 subjects (31.4%) had CSF CXCL13 above the limit of detection of the assay: median CSF CXCL13 level was 15.8 (8.2-91.0) pg/mL (Fig.1). Compared to subjects with undetectable CSF CXCL13, this group presented about two-fold higher CSF HIV-RNA, higher CSF proteins and neopterin levels, an increased CSAR levels (but no significantly larger proportion of BBB impairment), and higher values of Tourtelotte index, as shown in Tab.1. Interestingly, despite more subjects with detectable CSF CXCL13 had intrathecal synthesis (66.7%), the difference was not significant due to the presence of this phenomenon also in 49.1% of subjects with undetectable CXCL13 (Tab.1). At sensitivity analysis, after removing the subjects with altered BBB, those with detectable CSF CXCL13 (19/66, 28.8%; median CSF CXCL13 15.8 (8.2-48.0) pg/mL; Fig.1) confirmed to have higher CSF HIV-RNA, neopterin and proteins, as well as higher values of Tourtelotte index (see Suppl.Tab.1). No further demographic, clinical, viro-immunological parameters nor other CSF biomarkers differed between the two groups.

**Figure 1.**
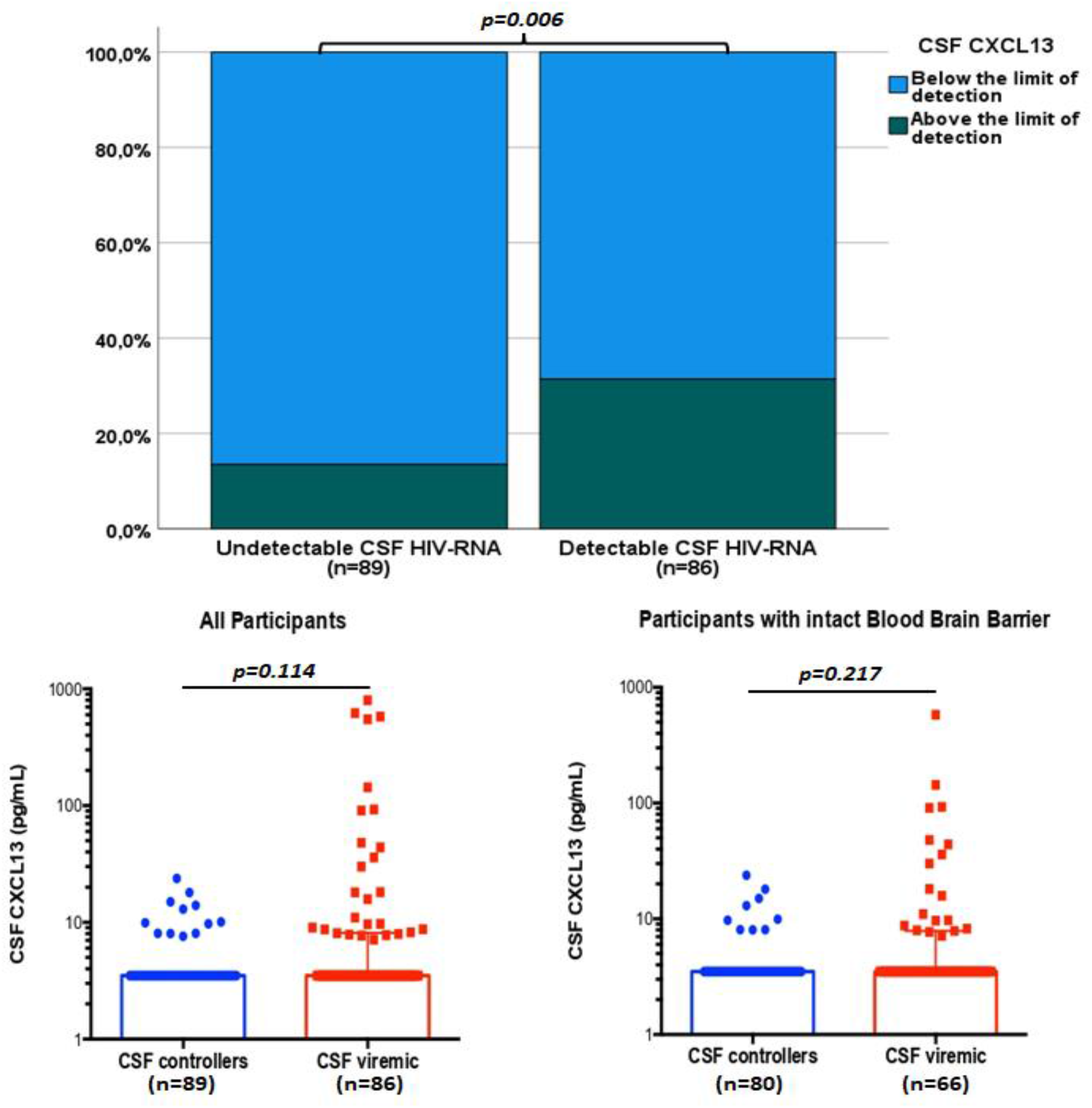
CSF CXCL13 detectability and levels in CSF viremic and controller subjects. CSF CXCL13 were Log10 transformed. Legend: CSF, Cerebrospinal fluid; BBB, Blood-brain barrier. Statistically significant comparisons were highlighted with the respective p-values.

CSF CXCL13 levels positively correlated with CSF HIV-RNA (rho 0.470, p=0.013; and not with plasma viremia), Tourtelotte (rho 0.404, p=0.037), CSAR (rho 0.724, p<0.001), IgG index (rho 0.711, p<0.001), with the proportion of immunoglobulins of intrathecal origin over the whole amount of CSF immunoglobulins (CSF % synthesis, rho 0.286, p=0.046), CSF S100beta and neopterin levels (rho 0.489, p=0.015 and 0.803, p<0.001, respectively), and with CSF proteins (rho 0.843, p<0.001). Considering the strong correlation between CSF CXCL13 and CSAR, despite the small sample, we run the correlations also among the 19 subjects without BBB impairment only. The majority of the previous findings was confirmed. Specifically, CSF CXCL13 did not correlate anymore with CSAR and CSF HIV-RNA, but its concentrations were associated with Tourtelotte (rho 0.832, p<0.001), Tibbling (rho 0.586, p=0.008), IgG index (rho 0.819, p<0.001), CSF % synthesis (rho 0.321, p=0.038), CSF neopterin (rho 0.735, p<0.001), and CSF cells and proteins (rho 0.790, p<0.001 and rho 0.788, p<0.001, respectively). We eventually tested the potential co-linearity of these correlations through bivariate linear regressions. Co-linearity was confirmed between CSF CXCL13 and CSF HIV-RNA, Tourtelotte, CSAR, IgG index, CSF % synthesis, CSF S100beta, CSF neopterin and CSF proteins, as shown in Tab.2.

**Table 2.** Bivariate linear regressions between CSF CXCL13 (pg/mL) and parameters with significant correlations in CSF viremic subjects. CSF, cerebrospinal fluid; CSAR, cerebrospinal fluid-to-serum albumin ratio.

|  | **B (95%CI)** | **P** |
| --- | --- | --- |
| **CSF HIV-RNA, Log10 cp/mL** | 76.3 (17.3-135.3) | **0.013** |
| **Tourtelotte** | 2.65 (0.18-5.13) | **0.037** |
| **Tibbling** | 17.8 (-140.9-176.4) | 0.820 |
| **CSAR** | 26.5 (16.1-36.9) | **<0.001** |
| **IgG index** | 356.6 (211.3-501.9) | **<0.001** |
| **CSF % synthesis, %** | 1.48 (0.024-2.94) | **0.046** |
| **CSF S100beta, ng/mL** | 0.99 (0.21-1.77) | **0.015** |
| **CSF neopterin, pg/mL** | 15.7 (10.6-20.9) | **<0.001** |
| **CSF proteins, mg/dL** | 4.72 (3.4-5.96) | **<0.001** |
| **CSF cells, cells/mm<sup>3</sup>** | 6.24 (-3.06-15.55) | 0.179 |

### 2. CSF CXCL13 in CSF controllers

Eighty-nine subjects had undetectable CSF HIV-RNA, of whom 17 (19.1%) with still detectable plasma viremia (median plasma HIV-RNA among them 63 (38-124) cp/mL). They were mostly Caucasian (91.0%) male (69.7%), with a long history of infection (median 10.4 years), valid immunological status (median CD4+T cell count and CD4/CD8 ratio of 475 (336-706) cells/mmc and 0.8 (0.5-1.2)) and presenting undetectable plasma viremia since a median time of 2.2 years, as shown in Tab.3. The prevalence of BBB impairment and intrathecal synthesis were 10.1% and 28.1%, respectively Twelve subjects (13.5%) had detectable CSF CXCL13: median CSF CXCL13 level was 10.0 (8.1-14.2) pg/mL and it did not differ significantly compared to the median CSF CXCL13 level in CSF viremic (p=0.114; Fig.1); on the contrary, the proportion of subjects with CSF CXCL13 above the limit of detection of the assay was significantly larger in the presence of CSF replicating virus compared to that of CSF controllers (31.4% vs 13.5%, OR 2.9 (1.4-6.3), p=0.006; Fig.1).

**Table 3.** Comparison between CSF controllers with versus without detectable CSF CXCL13: demographic, clinical, viro-immunological and CSF biomarkers. CXCL13, C-X-C motif chemokine ligand 13; MSM, males who have sex with other males; pIVDU, previous intra-venous drug users; PI, protease inhibitor; NRTIs, nucleoside reverse transcriptase inhibitors; nNRTI, non-nucleoside reverse transcriptase inhibitor; INI, integrase strand transfer inhibitor; CSF, cerebrospinal fluid; HAND, HIV-Associated neurocognitive disorders; CNS, central nervous system; CSAR, cerebrospinal fluid-to-serum albumin ratio; BBB, blood-brain barrier; S100b, S100 beta protein.

| Parameter | Subjects with undetectable CSF HIV-RNA (n=89) | Detectable CSF CXCL13 (n=12) | Undetectable CSF CXCL13 (n=77) | P |
| --- | --- | --- | --- | --- |
| Age, years | 51 (44-57) | 54 (45-68) | 50 (44-57) | 0.600 |
| Male sex, n | 62 (69.7%) | 8 (66.7%) | 54 (70.1%) | 0.808 |
| Caucasian, n | 81 (91.0%) | 12 (100%) | 69 (89.6%) | 0.242 |
| Exposure route, n |  |  |  | 0.310 |
| MSM | 39 (43.8%) | 5 (41.7%) | 34 (44.2%) |  |
| Heterosexual | 20 (22.5%) | 1 (8.3%) | 19 (24.7%) |  |
| pIVDU | 30 (33.7%) | 6 (50.0%) | 24 (31.2%) |  |
| Regimen, n |  |  |  | 0.562 |
| Dual | 15 (16.8%) | 4 (33.3%) | 11 (14.3%) |  |
| PI-2NRTIs | 31 (34.8%) | 3 (25.0%) | 28 (36.4%) |  |
| nN-2NRTIs | 13 (14.6%) | 2 (16.7%) | 11 (14.3%) |  |
| INI-2NRTIs | 20 (22.5%) | 2 (16.7%) | 18 (23.4%) |  |
| Others | 10 (11.2%) | 1 (8.3%) | 9 (11.7%) |  |
| Plasma HIV-RNA, Log10 cp/mL | 1.28 (0.04-1.28) | 1.28 (0.04-1.28) | 1.28 (0.04-1.28) | 0.558 |
| Detectable plasma HIV-RNA, n | 17 (19.1%) | 0 (0.0%) | 17 (22.1%) | 0.070 |
| CD4 count, cells/mm<sup>3</sup> | 475 (336-706) | 560 (394-920) | 466 (318-701) | **0.019** |
| CD4/CD8 ratio | 0.8 (0.5-1.2) | 1.2 (0.7-1.9) | 0.7 (0.5-1.1) | **0.040** |
| CD4 nadir, cells/mm<sup>3</sup> | 177 (63-263) | 201 (48-237) | 154 (63-263) | 0.509 |
| Length of HIV infection, months | 125 (38-225) | 59 (38-231) | 128 (38-225) | 0.793 |
| Length of plasma suppression, months | 26 (8-76) | 29 (9-49) | 26 (8-87) | 0.144 |
| Time on last cART regimen, months | 12 (8-34) | 10 (11-46) | 12 (7-25) | 0.174 |
| Clinical category, n |  |  |  | **0.008** |
| Neurological complaints | 22 (24.7%) | 2 (16.7%) | 20 (25.9%) | 0.487 |
| HAND | 29 (32.6%) | 7 (58.3%) | 22 (28.6%) | **0.041** |
| Asymptomatic | 37 (41.6%) | 2 (16.7%) | 35 (45.4%) | **0.059** |
| CNS HIV-related | 1 (1.1%) | 1 (8.3%) | 0 (0.0%) | **0.011** |
| Previous syphilis, n | 18 (20.2%) | 4 (33.3%) | 14 (18.2%) | 0.224 |
| CSF cells | 0 (0-0) | 0 (0-0) | 0 (0-0) | 0.651 |
| CSF proteins | 40 (31-49) | 50 (38-58) | 39 (30-48) | **0.007** |
| CSF glucose | 58 (54-64) | 63 (54-74) | 58 (54-63) | 0.066 |
| Tourtelotte | 0.0 (0.0-6.4) | 1.9 (0.0-10.1) | 0.0 (0.0-5.0) | **0.027** |
| Tibbling | 0.6 (0.4-0.8) | 0.8 (0.4-1.4) | 0.6 (0.4-0.7) | **0.054** |
| CSAR | 4.8 (3.8-6.3) | 5.5 (3.9-6.2) | 4.7 (3.5-6.1) | 0.135 |
| Impaired BBB, n | 9 (10.1%) | 3 (25.0%) | 6 (7.8%) | 0.065 |
| IgG index | 0.30 (0.21-0.45) | 0.37 (0.25-0.64) | 0.29 (0.19-0.43) | **0.021** |
| CSF synthesis, n | 25 (28.1%) | 7 (58.3%) | 18 (23.4%) | **0.012** |
| CSF % synthesis, % | 34.0 (17.5-51.5) | 42.0 (12.0-61.0) | 31.0 (19.0-50.7) | 0.612 |
| **Neopterin** | 0.6 (0.4-1.2) | 1.4 (0.6-2.5) | 0.56 (0.36-1.0) | **<0.001** |
| **S100b** | 110.2 (77.6-184.6) | 136.0 (91.4-207.9) | 110.0 (69.1-184.6) | 0.088 |

Among CSF controllers, those with detectable CSF CXCL13 presented higher CD4+T cell count and CD4/CD8 ratio, but also higher proportion of HAND and of HIV-related CNS disorders (Fig.2), and higher levels of CSF proteins, neopterin, and of biomarkers of intrathecal synthesis compared to those with undetectable chemokine, as shown in Tab.3. At sensitivity analysis, after removing the subjects with altered BBB, those with detectable CSF CXCL13 (9/80, 11.2%; median CSF CXCL13 of 9.9 (8.0-16.5) pg/mL) confirmed to have higher CD4+T cell count and CD4/CD8 ratio, a higher proportion of HAND (Fig.2), and the same pattern of CSF biomarkers (see Suppl.Tab.2). No further demographic, clinical, viro-immunological parameters nor other CSF biomarkers differed between the two groups.

**Figure 2.**
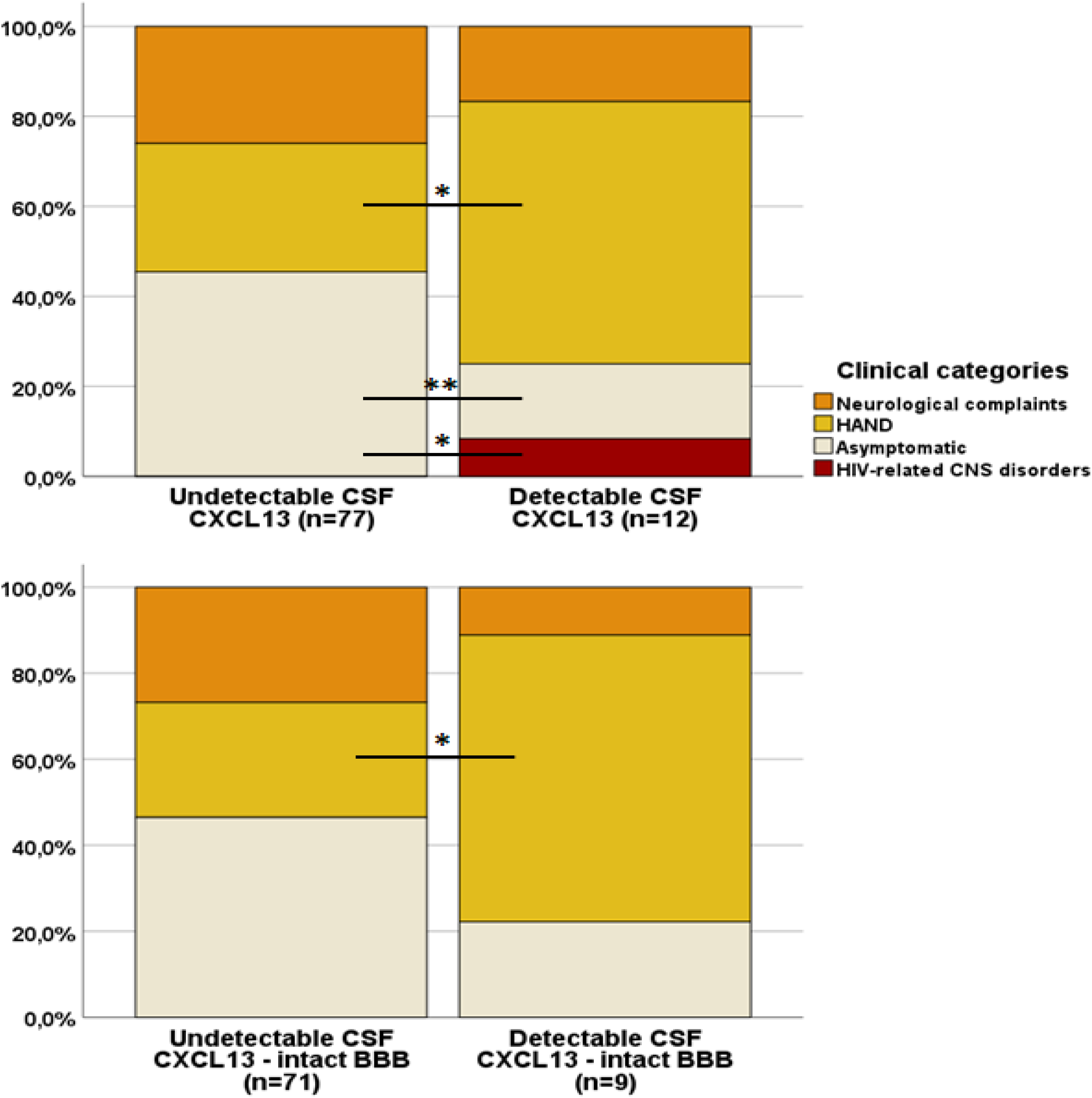
Comparison of the distribution of the neurological clinical categories among CSF controllers with versus without detectable CSF CXCL13. Legend: HAND, HIV-Associated neurocognitive disorders; CNS, Central Nervous system. Statistically significant differences were highlighted: * p<0.05, and **p=0.059.

CSF CXCL13 levels positively correlated with CD4+ T cell count (rho 0.763, p=0.006), CD4+ T cell nadir (rho 0.583, p=0.047), and CSF cells (rho 0.745, p=0.005). Despite the extremely limited sample (n=9), the same correlations were confirmed among subjects with no BBB impairment: CD4+ T cell count (rho 0.880, p=0.004), CD4+ T cell nadir (rho 0.688, p=0.041) and CSF cells (rho 0.768, p=0.016). We eventually tested the potential co-linearity of these correlations through bivariate linear regressions. Co-linearity was confirmed between CSF CXCL13 and CD4+ T cell count (B 0.009 (95%CI: 0.003-0.015), p=0.006) and CD4+ T cell nadir (B 0.015 (95%CI: 0.0005-0.029), p=0.047), but not with CSF cells (B 4.0 (95%CI: -2.4-10.4), p=0.503).

## Discussion

We described for the first time the presence and levels of CSF CXCL13 among cART-naive PLWH in advanced stage of infection (31.4%) and among cART-treated PLWH with a long history of previously untreated infection but substantial immunological recovery after two years of antiretroviral treatment and undetectable CSF HIV-RNA (13.5%). Only two previous studies reported CSF CXCL13 in PLWH without evidence of concurrent CNS infections: Bremell et al. detected 51.8% CSF samples with CXCL13 above the limit of detection of the assay among 27 asymptomatic cART-naive subjects (Bremell et al. 2013). The median CSF CXCL13 level was 10 pg/mL, which was very similar to the median level we measured in our cART-naive subjects, despite a different ELISA assay, higher CD4+T cell count, and lower median CSF HIV-RNA compared to ours (Bremell et al. 2013). Van Burgel reported on 7 HIV-positive subjects with no neurological complaints, finding extremely low CSF CXCL13 levels (median 1 pg/mL)(van Burgel et al. 2011). The authors concluded that CSF CXCL13 increases only during concomitant CNS infections or HIV encephalitis/meningitis, as compared to the increased levels they observed in HIV-positive subjects with opportunistic, non-opportunistic and HIV-mediated CNS infections (van Burgel et al. 2011) and as described by studies on neurosyphilis in PLWH (Marra et al. 2010). While Bremell could not entirely rule out concomitant subclinical CNS infections, we extensively assessed our patients excluding even the lowest CSF replication of CMV and EBV, that have been previously associated with BBB impairment and increased intrathecal synthesis (Bonnan et al. 2015; Caligaris et al. 2021). Therefore, our data do not support the necessity of other infections nor of clinically relevant HIV infection of the CNS for the intrathecal production of CXCL13. This is also expected considering that human monocytes, which are abundant in CNS, have been proved to be able to produce CXCL13 after direct exposure to HIV-1 single strand RNA (Cohen et al. 2015).

Among cART-naive subjects, the amount of CSF virus directly correlated with the amount of measured CSF CXCL13 demonstrating the strongest increase in CXCL13 per Log10 unit more of HIV-RNA (after IgG index, that is directly linked to intrathecal B and plasma cells activation). It is interesting to note that even in the absence of replicating virus, still 13.5% of PLWH presented detectable CSF CXCL13. Therefore, while our data confirm that replicating HIV-RNA is an important stimulus to CXCL13 production even within CNS, it is not necessary an overt viral replication to induce the production of this chemokine, as already observed for plasma CXCL13 (Widney et al. 2005; Regidor et al. 2011; Mehraj et al. 2019). Unfortunately, we were not able to collect any data on CNS reservoir, and due to the invasiveness of CSF collection, we were not able to include an HIV-negative healthy control group to compare prevalence and levels of CSF CXCL13.

Conversely, despite actively replicating virus, 68.6% of cART-naive subjects had no detectable CSF CXCL13. This finding should take into account potential measurement bias, as we did not perform duplicates measurements and some samples remained frozen for up to 10 years before the analysis. Indeed, the cell source of CXCL13 within CNS has not been identified yet, the daily recirculation of CXCL13 levels within CSF is unknown (partially limited by performing all the LP in 4-hours span during the morning shift), and CXCL13 polymorphisms may affect CXCL13 levels (Hussain et al. 2013). All these considerations may explain the high prevalence of negative CSF CXCL13 despite local viral replication.

Compared to previously reported values of plasma CXCL13 (Widney et al. 2005; Regidor et al. 2011; Mehraj et al. 2019), the CSF levels we detected were significantly lower (of about 1 Log10) both when considering plasma concentrations in cART-naive and in cART-treated PLWH. Indeed, cART seems to reduce but not normalize plasma CXCL13 after up to 2-3 years of follow up (Widney et al. 2005; Regidor et al. 2011; Mehraj et al. 2019). The main hypothesis on the persistently increased level of plasma CXCL13 despite viral suppression relies on the potential chronic stimulation of CXCL13-producing cells by HIV proteins within secondary lymphoid tissues. While the observed strength of the correlation between CSF CXCL13 and CSF neopterin, marker of monocyte/macrophages activation, may support the idea that the main cell source of this chemokine within CNS is represented by monocytes/macrophages, tailored studies should properly address this issue, identifying the cell lines involved in the CNS CXCL13/CXCR5 axis, and clarifying why some subjects do not produce intrathecal CXCL13 despite the presence of an acknowledged trigger such as HIV. This could also help at identifying specific CNS targets for functional cure strategies as it is for peripheral compartments. Recently a subset of CXCR5+ NK located in B cell follicles has been identified (Guo et al. 2022). Their frequency correlated with CXCL13 intensity in lymph nodes of treated PLWH, and, interestingly, an inverse relationship between this NK subset and HIV-DNA/RNA within the follicles was observed, suggesting that these cells may exert an inhibitory activity on HIV reservoir (Guo et al. 2022). Considering our finding and the brain cytoarchitecture, it is likely that the cell sources and the mechanisms underlying CXCL13 production within CNS differ from what has been observed is the periphery. Therapeutic CXCL13 blockade through monoclonal antibodies in animal models of autoimmune diseases has been associated with no evident toxicity, but with outcomes that variably changed from poor to clinically relevant benefit (Henry and Kendall 2010; Wu et al. 2015).

Another interesting finding consisted in the fact that, despite the presence of CSF CXCL13 was associated with more common and prominent intrathecal synthesis (as median IgG index and as daily intrathecal immunoglobulins synthesis expressed by Tourtelotte index), intrathecal synthesis was recorded even in cART-naive and cART-treated subjects with no detectable CSF CXCL13 (49.1% and 23.4%, respectively). In these cases, the proportion of immunoglobulins directly originating within the CNS over the overall amount of CSF immunoglobulins was also similar to the proportion measured in subjects with CSF CXCL13. B-cell trafficking into CNS and CNS-resident IgG-secreting cells are poorly characterized; most of the intrathecal IgG synthesis associated with the immune response to CNS viral infections appears to be non-specific (up to 95% of the intrathecally produced IgG), including HIV infection (Bonnan et al. 2015). Considering that CSF plasma cells seem to disappear after cART initiation while CSF IgG synthesis is mostly preserved (Bonnan et al. 2015), IgG-secreting cells are probably resident within CNS, but it is still unknown whether these cells are blood-borne or they mature inside CNS. As we detected intrathecal synthesis even in the absence of CXCL13, which is a B-cells chemoattractant chemokine, it is more likely that plasma cells and IgG-secreting cells have been primed, matured and induced to IgG secretion directly within CNS. Further studies are warranted to identify also potential alternative pathways involved in CNS homing and CSF recirculation of IgG-secreting cells that do not require the activation of CXCL13/CXCR5 axis.

The relationship between plasma CXCL13 and CD4+T cell count, CD4/CD8 ratio and CD4+T cells nadir is debated, as the plasma levels of this chemokine have been described to be both positively and negatively correlated with CD4+T cells (Widney et al. 2005; Mehraj et al. 2019). We observed that CSF controllers with detectable CSF CXCL13 had higher CD4+T cells count and higher CD4/CD8 ratio; our data also suggest that the value of CXCL13 changed according to a positive co-linear relationship with current and nadir CD4+ T cell count. The interpretations of this finding could be multiple. The association could be a sign of a better immunological functioning, either as higher absolute CD4+T cells values, more balanced ratio among T cell subtypes, and chronic humoral responses and immuno surveillance perhaps involved in viral control together with antiretroviral treatment. Nevertheless the persistence and increased plasma levels of CXCL13 have been associated with higher mortality and correlated with other biomarkers of detrimental processes in PLWH, such as microbial translocation and immune activation (Wada et al. 2016; Mehraj et al. 2019). Furthermore, higher values of CD4+ T cells per sé do not necessarily associate with better immunological status, as the relative subsets of circulating CD4+ T cells and their proportion can better represent functionality of this cell population. Indeed, CXCR5+ circulating Tfh have been reported to be associated with CXCL13 levels and to expand after cART initiation (Niessl et al. 2020); despite HIV-specific, this large subset of CD4+ T cells possesses multiple phenotypic and functional features less prone to viral suppression than non-circulating HIV-specific Tfh and have been correlated with the amount of translation-competent HIV reservoir (Niessl et al. 2020). Interestingly, increased presence of CXCR5+CD4+ T cells along with intrathecal CXCL13 elevation has been recently described in a broad spectrum of inflammatory CNS diseases, ranging from aseptic meningoencephalitis to CNS autoimmune disorders (Harrer et al. 2021). Therefore, another very hypothetical explanation of the observed relationship between the presence of CSF CXCL13 and increased CD4+ T cells count could be that this association simply detected the expansion of CXCR5+CD4+ T cells that could come along increased levels of the chemokine. It is noteworthy that in the study by Harrer et al., increased CSF CXCL13 associated with increased intrathecal CXCR5+CD4+ T cell frequencies and absolute number (Harrer et al. 2021). Translating this observation to HIV infection, CXCL13/CXCR5 axis could amplify (by a trojan horse mechanism) the number of cells harbouring HIV within the CNS, suggesting once more its negative role when chronically hyper-activated. CSF controllers with detectable CSF CXCL13 were also characterized by increased CSF levels of proteins, neopterin and intrathecal synthesis, all variables related with negative CNS outcomes (Bonnan et al. 2015; Motta et al. 2017; Barco et al. 2022). These subjects suffered more commonly from HAND compared to subjects with undetectable CSF CXCL13. One subject with detectable CSF CXCL13 versus none among those with undetectable chemokine had also persistence of HIV-associated dementia and spastic tetraparesis with a brain MRI imaging compatible with advanced HIV encephalopathy despite being on effective treatment since more than two years. In multiple sclerosis, which shows several overlaps with HIV-related CNS inflammatory processes (Bonnan et al. 2015), CSF CXCL13 has been usually reported as moderately higher (20 pg/mL up to more tha 500 pg/mL) compared to what we measured in our study population (Khademi et al. 2011; Alvarez et al. 2013; DiSano et al. 2020; Pitteri et al. 2022). Increased CSF CXCL13 levels have been linked with disease activity, relapse rate, number of brain lesions and unfavourable overall prognosis (Khademi et al. 2011; Alvarez et al. 2013; DiSano et al. 2020). Interestingly, CSF CXCL13 levels were used to discriminate mild from severe cognitive impairment in patients suffering from multiple sclerosis (Pitteri et al. 2022). In PLWH, increased intrathecal synthesis has been previously associated with a worse neurocognitive outcome (Bonnan et al. 2015; De Almeida et al. 2021); herein, we have introduced in this relationship a further piece represented by CSF CXCL13 that could be therefore regarded as a potential biomarker candidate for neurocognition and HAND in this population.

Our study presents different limitations: the small sample size, the retrospective design, the long conservation of frozen CSF samples that may have affected the exact quantification of CXCL13, and the large between-subjects variability in CSF CXCL13. Regarding this last issue, while we cannot rule out increased variability not limited by repeated measurements per sample, it could also be a sign of different physiological and pathological processes involving this chemokine in our sample. Larger studies should address this issue stratifying by CSF CXCL13 levels to discriminate potentially physiological and favourable levels from pathological concentrations.

In conclusion, the presence of detectable CXCL13 CSF concentrations could identify a subgroup of HIV-positive subjects presenting increased CNS IgG synthesis, chronic inflammation, and immune activation. Among subjects on antiretroviral therapy and undetectable CSF HIV-RNA, the presence of CXCL13 in CSF was also associated with increased likelihood of neurocognitive impairment and HIV-related CNS disorders. This hypothesis-generating work raises questions that will need to be addressed by future prospective studies to identify the cell source of CXCL13 within CNS and its potential roles as perpetrator of brain injury and inflammation, as diagnostic biomarker of HAND and as target for immune-based therapeutic strategies.

## Data Availability

All data produced in the present study are available upon reasonable request to the authors

