## Supplementary material for "Cerebrospinal fluid CXCL13 identifies a subgroup of people living with HIV with prominent intrathecal synthesis, immune activation, and neurocognitive impairment regardless of effective antiretroviral therapy": Suppl.Tab.

**Supplementary Table 1. Comparison between CSF viremic subjects and intact blood-brain barrier with versus without detectable CSF CXCL13: demographic, clinical, viro-immunological and CSF biomarkers**

| Parameter | Detectable CSF CXCL13 (n=19) | Undetectable CSF CXCL13 (n=47) | P |
|---|---|---|---|
| **Age, years** | 48 (38-57) | 46 (39-50) | 0.449 |
| **Male sex, n** | 15 (78.9%) | 34 (72.3%) | 0.578 |
| **Caucasian, n** | 18 (94.7%) | 38 (80.8%) | 0.154 |
| **Exposure route, n** | | | 0.617 |
| **MSM** | 7 (36.8%) | 13 (27.6%) | |
| **Heterosexual** | 7 (36.8%) | 16 (34.0%) | |
| **pIVDU** | 5 (26.3%) | 18 (38.3%) | |
| **On cART, n** | 7 (36.8%) | 18 (38.3%) | 0.912 |
| **Regimen, n** | | | 0.446 |
| **Dual** | 2/7 (28.6%) | 2/18 (11.1%) | |
| **PI-2NRTIs** | 0/7 (0%) | 6/18 (33.3%) | |
| **nN-2NRTIs** | 1/7 (14.3%) | 3/18 (16.7%) | |
| **INI-2NRTIs** | 1/7 (14.3%) | 2/18 (11.1%) | |
| **Others** | 3/7 (42.8%) | 5/18 (27.8%) | |
| **Plasma HIV-RNA, Log10 cp/mL** | 5.13 (1.60-5.83) | 5.18 (1.74-5.71) | 0.882 |
| **Undetectable plasma HIV-RNA, n** | 3 (15.8%) | 10 (21.2%) | 0.803 |
| **CSF HIV-RNA, Log10 cp/mL** | 4.17 (2.18-4.63) | 2.56 (2.10-3.70) | ***0.018*** |
| **CSF escape, n** | 4 (21.0%) | 9 (19.1%) | 0.861 |
| **CD4 count, cells/mmc** | 70 (33-194) | 83 (29-282) | 0.613 |
| **CD4/CD8 ratio** | 0.1 (0.1-0.2) | 0.2 (0.1-0.5) | 0.286 |
| **CD4 nadir, cells/mmc** | 48 (15-99) | 47 (16-141) | 0.502 |
| **Time since HIV diagnosis, months** | 79 (1-150) | 41 (1-180) | 0.810 |
| **Clinical category, n** | | | 0.943 |
| **Neurological complaints** | 2 (10.5%) | 5 (10.6%) | 0.968 |
| **HAND** | 4 (21.0%) | 7 (14.9%) | 0.416 |
| **Asymptomatic** | 8 (42.1%) | 21 (44.7%) | 0.849 |
| **HIV-related** | 5 (26.3%) | 14 (29.8%) | 0.740 |
| **Previous syphilis, n** | 1 (5.3%) | 8 (17.0%) | 0.207 |
| **CSF cells** | 0 (0-1) | 0 (0-0) | 0.980 |
| **CSF proteins** | 49 (35-56) | 39 (29-49) | ***0.039*** |
| **CSF glucose** | 51 (41-60) | 53 (49-61) | 0.260 |
| **Tourtelotte** | 16.4 (4.6-47.1) | 5.5 (0.3-13.6) | ***0.008*** |
| **Tibbling** | 1.0 (0.8-1.3) | 0.8 (0.6-1.1) | 0.062 |
| **CSAR** | 5.3 (3.4-6.3) | 4.4 (3.2-5.9) | 0.348 |
| **IgG index** | 0.55 (0.32-0.69) | 0.37 (0.24-0.50) | 0.616 |
| **CSF synthesis, n** | 15 (78.9%) | 25 (53.2%) | ***0.052*** |
| **CSF % synthesis, %** | 38.0 (30.5-50.0) | 36.0 (25.0-44.0) | 0.301 |
| **Neopterin** | 3.6 (0.9-9.2) | 1.1 (0.9-2.1) | ***<0.001*** |
| **S100b** | 136.9 (59.2-298.4) | 129.4 (89.2-173.2) | 0.704 |

Legend: CXCL13, C-X-C motif chemokine ligand 13; MSM, males who have sex with other males; pIVDU, previous intra-venous drug users; PI, protease inhibitor; NRTIs, nucleoside reverse transcriptase inhibitors; nNRTI, non-nucleoside reverse transcriptase inhibitor; INI, integrase strand transfer inhibitor; CSF, cerebrospinal fluid; HAND, HIV-Associated neurocognitive disorders; CNS, central nervous system; CSAR, cerebrospinal fluid-to-serum albumin ratio; S100b, S100 beta protein.

**Supplementary Table 2. Comparison between CSF controllers and intact blood-brain barrier with versus without detectable CSF CXCL13: demographic, clinical, viro-immunological and CSF biomarkers**

| Parameter | Detectable CSF CXCL13 (n=9) | Undetectable CSF CXCL13 (n=71) | P |
|---|---|---|---|
| **Age, years** | 54 (45-68) | 50 (44-56) | 0.172 |
| **Male sex, n** | 6 (66.7%) | 50 (70.4%) | 0.818 |
| **Caucasian, n** | 9 (100%) | 63 (88.7%) | 0.292 |
| **Exposure route, n** | | | 0.512 |
| **MSM** | 4 (44.4%) | 31 (43.7%) | |
| **Heterosexual** | 0 (0.0%) | 17 (23.9%) | |
| **pIVDU** | 5 (55.5%) | 23 (32.4%) | |
| **Regimen, n** | | | 0.902 |
| **Dual** | 2 (22.2%) | 10 (14.1%) | |
| **PI-2NRTIs** | 2 (22.2%) | 26 (36.6%) | |
| **nN-2NRTIs** | 2 (22.2%) | 11 (15.5%) | |
| **INI-2NRTIs** | 2 (22.2%) | 16 (22.5%) | |
| **Others** | 1 (11.1%) | 8 (11.3%) | |
| **Plasma HIV-RNA, Log10 cp/mL** | 1.28 (0.04-1.28) | 1.28 (0.04-1.28) | 0.688 |
| **Detectable plasma HIV-RNA, n** | 0 (0.0%) | 16 (22.5%) | 0.111 |
| **CD4 count, cells/mmc** | 560 (394-920) | 466 (313-704) | ***0.043*** |
| **CD4/CD8 ratio** | 1.2 (0.7-1.9) | 0.7 (0.4-1.1) | ***0.018*** |
| **CD4 nadir, cells/mmc** | 201 (49-238) | 153 (50-253) | 0.615 |
| **Length of HIV infection, months** | 59 (38-231) | 135 (39-222) | 0.814 |
| **Length of plasma suppression, months** | 29 (9-49) | 26 (8-89) | 0.265 |
| **Time on last cART regimen, months** | 9 (9-41) | 12 (6-26) | 0.142 |
| **Clinical category, n** | | | ***0.052*** |
| **Neurological complaints** | 1 (11.1%) | 19 (26.8%) | 0.307 |
| **HAND** | 6 (66.7%) | 19 (26.8%) | ***0.015*** |
| **Asymptomatic** | 2 (22.2%) | 33 (46.5%) | 0.167 |
| **HIV-related** | 0 (0.0%) | 0 (0.0%) | - |
| **Previous syphilis, n** | 3 (33.3%) | 14 (19.7%) | 0.350 |
| **CSF cells** | 0 (0-0) | 0 (0-0) | 0.758 |
| **CSF proteins** | 50 (38-58) | 39 (29-47) | ***0.034*** |
| **CSF glucose** | 63 (54-74) | 58 (54-62) | 0.131 |
| **Tourtelotte** | 2.9 (0.0-11.9) | 0.0 (0.0-6.2) | ***0.027*** |
| **Tibbling** | 0.8 (0.4-1.4) | 0.6 (0.5-0.7) | ***0.023*** |
| **CSAR** | 5.5 (3.9-6.2) | 4.5 (3.2-5.8) | 0.258 |
| **IgG index** | 0.37 (0.25-0.64) | 0.28 (0.19-0.39) | ***0.023*** |
| **CSF synthesis, n** | 6 (66.7%) | 18 (25.3%) | ***0.011*** |
| **CSF % synthesis, %** | 42.0 (12.0-61.0) | 31.0 (19.0-50.7) | 0.612 |
| **Neopterin** | 1.4 (0.6-2.5) | 0.6 (0.4-1.0) | ***0.001*** |
| **S100b** | 136.0 (91.4-207.9) | 110.2 (69.2-183.4) | 0.916 |

Legend: CXCL13, C-X-C motif chemokine ligand 13; MSM, males who have sex with other males; pIVDU, previous intra-venous drug users; PI, protease inhibitor; NRTIs, nucleoside reverse transcriptase inhibitors; nNRTI, non-nucleoside reverse transcriptase inhibitor; INI, integrase strand transfer inhibitor; CSF, cerebrospinal fluid; HAND, HIV-Associated neurocognitive disorders; CNS, central nervous system; CSAR, cerebrospinal fluid-to-serum albumin ratio; S100b, S100 beta protein.
